# A Decade of Progress: Description of Ebola Vaccination Strategy and its Associated Factors in Sierra Leone

**DOI:** 10.64898/2026.01.28.26345081

**Authors:** Desmond M. Kangbai, Edwin N. Sesay, Nella Clemens-Kangbai, Hassan Benya, Peter Bai James, Henry Carter, Andrew K. Kemoh, Sallu Lansana, Edna G. Samuels, Mohamed T. M. Kallay, Nelson Mandela Kargbo, Jia Kangbai, Francis Moses

**Affiliations:** Expanded Program on Immunisation, Ministry of Health and Sanitation; College of Medicine and Allied Health Sciences, University of Sierra Leone; Clinton Health Access Initiative, Sierra Leone; Department of Public Health, School of Public Health, Njala University, Sierra Leone; Directorate of Reproductive and Maternal Health, Ministry of Health, Sierra Leone

**Author notes:** **Correspondence:** Desmond Maada Kangbai, Central Medical Stores, New England Ville.

**Keywords:** Ebola, vaccines, community engagement, Sierra Leone

## Abstract

**Background:** The 2013-2016 Ebola virus disease outbreak in West Africa profoundly affected Sierra Leone’s health financing, health delivery, health governance, and health workforce. The country becomes the first among those most severely affected ten years ago by the West Africa Ebola outbreak, to launch nationwide preventive Ebola vaccination, targeting 20,000 frontline workers who received a single dose of the Ebola Vaccine.

**Methods:** A mixed-method study design that analysed administrative vaccination data, field supervision reports, and electronic records from all 16 districts in Sierra Leone was used. The following indicators were analysed: vaccination coverage and logistical regression. Qualitative information from stakeholder debriefings and community feedback complemented the findings from the quantitative analysis. Quantitative data were analysed using Statistical Package for the Social Sciences (SPSS) version 27.

**Results:** A total of 17,454 (vaccine coverage= 84.5%) frontline workers and high-risk populations for EVD infection were vaccinated. Healthcare workers accounted for 35% of all vaccinations. Multivariate analysis showed that females had an AOR of 1.507 (95% CI: 1.293-1.756, P < 0.001) and individuals with tertiary education had an AOR of 1.900 (95% CI: 1.371-2.633, P < 0.001), were more likely to be vaccinated. Western and Southern regions achieved the highest coverage due to superior cold-chain readiness and community mobilisation. No serious adverse events following immunisation were reported.

**Conclusion:** Sierra Leone’s decade-long journey from outbreak response to preventive vaccination demonstrates the transformative impact of sustained investment, multisectoral coordination, and community trust. High coverage and operational success affirm the feasibility of nationwide Ebola immunisation in resource-limited settings.

## Introduction

The Ebola Virus Disease (EVD) outbreak in Guinea, Liberia, and Sierra Leone, all in West Africa in 2013, was considered one of the most devastating EVD outbreaks since the Ebola virus was discovered in 1976 (1). The outbreak posed a significant threat to public health due to its high case-fatality rates, occurrences, and overall severity (2,3). In May 2014, Sierra Leone reported its first EVD cases in the eastern districts of Kailahun and Kenema. The outbreak then spread rapidly in the early stages across all districts in the country (4). The overall case-fatality ratio (CFR) estimates for the 2013-2016 West African Ebola epidemic were 82.2%. With Sierra Leone recording the highest CFR 89.1%, Liberia 79.2% and Guinea 65.6% respectively (5). Sierra Leone recorded the highest EVD mortality rate in 2014 ranged from 4468 to 15824, Liberia from 2928 to 10374 and Guinea from 1739 to 5548, respectively(6). The majority of those infected by the Ebola virus during the 2014-2016 outbreak in Sierra Leone were front-line healthcare workers. The outbreak also exposed the country’s already fragile healthcare infrastructure to near collapse (7). The outbreak revealed the severe effects of a lack of preparedness by the EVD repercussions of inadequately equipped health systems for addressing new infectious diseases (8).

According to the World Health Organisation (WHO), statistics show that the EVD outbreak in West Africa has emerged as the most devastating in history, possessing the capacity to instil global fear, frequently exacerbated by migrant Ebola cases and sensationalist media coverage (9). The high EVD Case fatality rate and morbidity rates during the 2013-2016 EVD outbreak provide a perfect timing for designing and experimentation of an EVD vaccine. During and even after the 2013-2016 EVD outbreak, there were tremendous strides from global manufacturers to develop Ebola vaccines (10). Vaccination has since emerged as a critical component of EVD prevention and control (11). The rapid development and deployment of Ebola vaccines during the outbreak on compassionate grounds highlighted the crucial role of immunisation in mitigating public health emergencies. This effort established the foundation for the development and implementation of long-term strategies to prevent future outbreaks and enhance global health preparedness (12).

A heterologous two-dose vaccination regimen with adenovirus 26 (Ad26) and modified vaccinia virus Ankara 85 (MVA) expressing the glycoprotein of Zaire Ebola virus (Ad26.ZEBOV and MVA-BN-Filo) was evaluated in an 86-randomised controlled trial in a community affected by Ebola during the West Africa epidemic. This vaccine 87 regimen has been shown to have an acceptable safety profile and to induce robust humoral immune responses in adults (13). The transition of Sierra Leone’s health system from post-conflict recovery to epidemic resilience has seen various policy changes in vaccination delivery. However, there is a notable absence of empirical evidence that critically describes the trajectory of these vaccination strategies alongside their associated determinants over the last decade. Without this decade-long perspective, policymakers lack the historical evidence base required to refine current strategies and address deep-seated factors influencing vaccine uptake.

## Methods

### Study Design and Population

A mixed-method study that analysed Ebola vaccination data and described the vaccination strategies that were initially implemented during the 2014-2016 EVD outbreak and subsequently in 2024. The study population whose vaccination data were analyzed include healthcare workers, traditional healers, social workers, motorcyclists, and security forces in Sierra Leone during the study period, with a focus on the population data and vaccination records available within the public health system.

### Study Setting and Period

Sierra Leone, a resource-constrained country in West Africa with a population of over 8 million people (14), was one of the epicentres of the 2014-2016 Ebola outbreak. The vaccination campaign took place across all 16 districts in the country.

## Data Analysis

### Data Sources and Collection

Quantitative data were retrieved from the Ebola vaccination registers for case-based data on Ebola vaccination status (date of vaccination and location), and from population-based surveys. Qualitative data were retrieved from field supervision reports, stakeholder debriefings and community feedback.

### Data Assurance and Analysis

Before data analysis, data cleaning and validation procedures were performed to ensure data entered was assessed for accuracy and completeness. Data was analysed using Statistical Package for Social Scientists version 27 to analyse descriptive statistics, which includes frequencies and percentages, for vaccination status, prevalence and sociodemographic characteristics. A bivariate logistic regression was used to examine the association between vaccination status and individual sociodemographic factors. Also, multivariate logistic regression was used to identify the independent sociodemographic determinants of Ebola vaccination status, adjusting for potential confounders. Odds ratios (ORs) and 95% confidence intervals (CIs) were calculated to quantify the strength and direction of the associations. Vaccination coverage for every district was calculated.

### Variables and Measurements

**Ebola Vaccination Status**: The primary outcome variable, categorised as “Previously vaccinated” or “Currently vaccinated” based on vaccination records.

**Prevalence of Vaccination**: This was calculated as the proportion of the population vaccinated at any point during the study period.

**Sociodemographic Determinants**: Potential independent variables that may influence vaccination status, including: Age, categorised into appropriate age groups; Sex (male, female); Region of residence; Education level.

### Target Population and Vaccination Coverage

The number of healthcare workers, traditional Healers social workers, motorcyclists, and security forces targeted in the Ebola vaccination campaign in Sierra Leone was informed by a detailed microplanning at the district level, which involves target groups enumeration; this leads to the identification of eligible recipients. For Sierra Leone, Ebola vaccination is estimated at 95% for herd immunity. This means that at least 95% of the targeted groups aged 18 and above need to be immunised.

Total population of target groups aged 18 and above for the Ebola vaccination in Sierra Leone = 20,621

Number of Vaccinees who received the Ebola vaccine = **17,454**

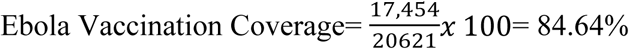

The target Ebola Vaccination coverage to achieve herd immunity= 20,621x 95%=**19,590** Total population of targeted healthcare workers = 7,780

Number of Vaccinees who received the Ebola Vaccine= 6,058

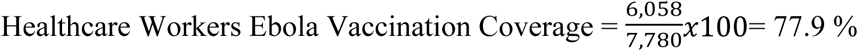

Total population of targeted Traditional Healers = 2,968 Number of Vaccinees who received the Ebola Vaccine= 1,219

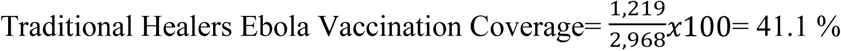

Number of Vaccinees who received the Ebola Vaccine= 3,051

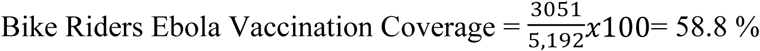

Total population of targeted Social Workers = 610

Number of Vaccinees who received the Ebola Vaccine=1,816

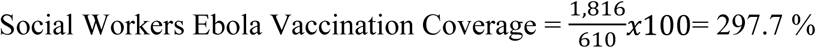

Total population of targeted Security Forces (Military, Police, Customs and Immigration Officers = 7,535

Number of Security Forces (Military, Police, Customs and Immigration Officers who received the Ebola Vaccine=5,312

Security Forces (Military, Police, Customs and Immigration Officers) Ebola Vaccination

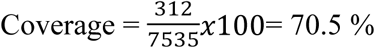

**Eligibility Criteria:** Targeted groups (healthcare workers, bike riders, security forces, traditional healers, and social workers for the Ebola vaccination who are age 18 and above and who are not immunocompromised are included in the study. Individuals who did not receive the Ebola vaccine at the time of vaccination are not included in the study.

**Ethical considerations:** Permission to conduct the study was granted by the Expanded Programme on Immunisation, through the Ministry of Health, Sierra Leone. Data were de-identified, stored on encrypted servers, and accessed only by authorised personnel. Participants were informed about voluntary participation and their right to withdraw without penalty.

## RESULTS

### Sociodemographic Characteristics of Ebola Vaccine Vaccinees

A total of 17,454 (84.5%) Ebola vaccine vaccinees were reported in the case-based register. The majority, 5,526 (31.7%) of the Ebola vaccine vaccinees belong to the age group 25-34 years. Regarding gender, males accounted for 11729 (67.2%). The study indicated that the 17,454 recipients of the Ebola vaccine, the majority 4,410 (25.3%), reside in the northern region of Sierra Leone. Furthermore, the majority, 11,978 (68.6%) of the recipients were married, and regarding educational level, the majority, 7001 (40.1%) of the recipients attained tertiary education, followed by secondary education, 6,259 (35.9%). In terms of occupation, the majority, 6,058 (35.9%) of the recipients were healthcare workers.

**Table 1:**
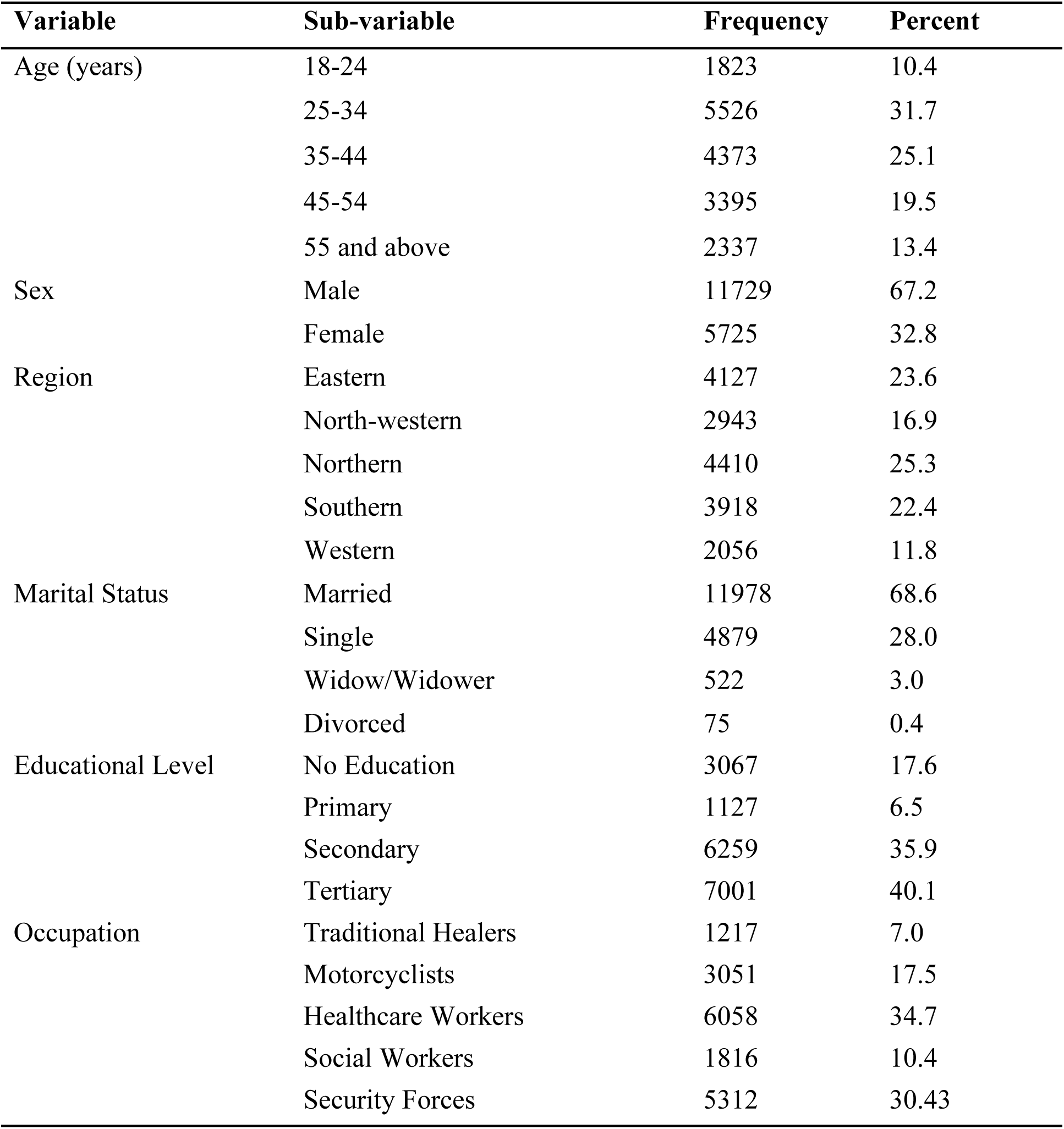
Socio-demographic Characteristics of Ebola Vaccine Vaccinees in Sierra Leone, December 2024 (N=17454)

| Variable | Sub-variable | Frequency | Percent |
| --- | --- | --- | --- |
| Age (years) | 18-24 | 1823 | 10.4 |
|  | 25-34 | 5526 | 31.7 |
|  | 35-44 | 4373 | 25.1 |
|  | 45-54 | 3395 | 19.5 |
|  | 55 and above | 2337 | 13.4 |
| Sex | Male | 11729 | 67.2 |
|  | Female | 5725 | 32.8 |
| Region | Eastern | 4127 | 23.6 |
|  | North-western | 2943 | 16.9 |
|  | Northern | 4410 | 25.3 |
|  | Southern | 3918 | 22.4 |
|  | Western | 2056 | 11.8 |
| Marital Status | Married | 11978 | 68.6 |
|  | Single | 4879 | 28.0 |
|  | Widow/Widower | 522 | 3.0 |
|  | Divorced | 75 | 0.4 |
| Educational Level | No Education | 3067 | 17.6 |
|  | Primary | 1127 | 6.5 |
|  | Secondary | 6259 | 35.9 |
|  | Tertiary | 7001 | 40.1 |
| Occupation | Traditional Healers | 1217 | 7.0 |
|  | Motorcyclists | 3051 | 17.5 |
|  | Healthcare Workers | 6058 | 34.7 |
|  | Social Workers | 1816 | 10.4 |
|  | Security Forces | 5312 | 30.43 |

### Clinical Characteristics of Ebola Vaccine Vaccinees in Sierra Leone

In terms of clinical characteristics of Ebola vaccine vaccinees, the majority, 17,243 (98.8%) of the vaccinees did not have a history of Ebola virus disease. In terms of allergies from previous vaccination, the majority, 17,385 (99.6%), did not experience any. Regarding co-morbidity, the majority, 17,306 (99.2%), did not have any. The majority of the Ebola vaccine vaccinees, 17,450 (99.9%), did not encounter Adverse Event Following Immunisation (AEFI).

**Table 2:** Clinical characteristics of Ebola Vaccine Recipients in Sierra Leone, December 2024 (N=17454)

| Variable | Sub-variable | Frequency | Percent |
| --- | --- | --- | --- |
| History of Ebola Virus Disease | Yes | 211 | 1.2 |
|  | No | 17243 | 98.8 |
| Allergies from Previous Vaccination | Yes | 69 | 0.4 |
|  | No | 17385 | 99.6 |
| Do you have any comorbidities | Yes | 148 | 0.8 |
|  | No | 17306 | 99.2 |
| AEFI Encountered | Yes | 4 | 0.1 |
|  | No | 17450 | 99.9 |

### Ebola Vaccination Status of Ebola Vaccine Vaccinees

The majority (n= 15,566, 89.2%) of the Ebola vaccine vaccinees were first-time vaccinees of the rVSV-ZEBOV Ebola vaccine; few (n = 1,888, 10.8%) vaccinees had received the Johnson & Johnson Ebola vaccine previously.

**Figure 1:**
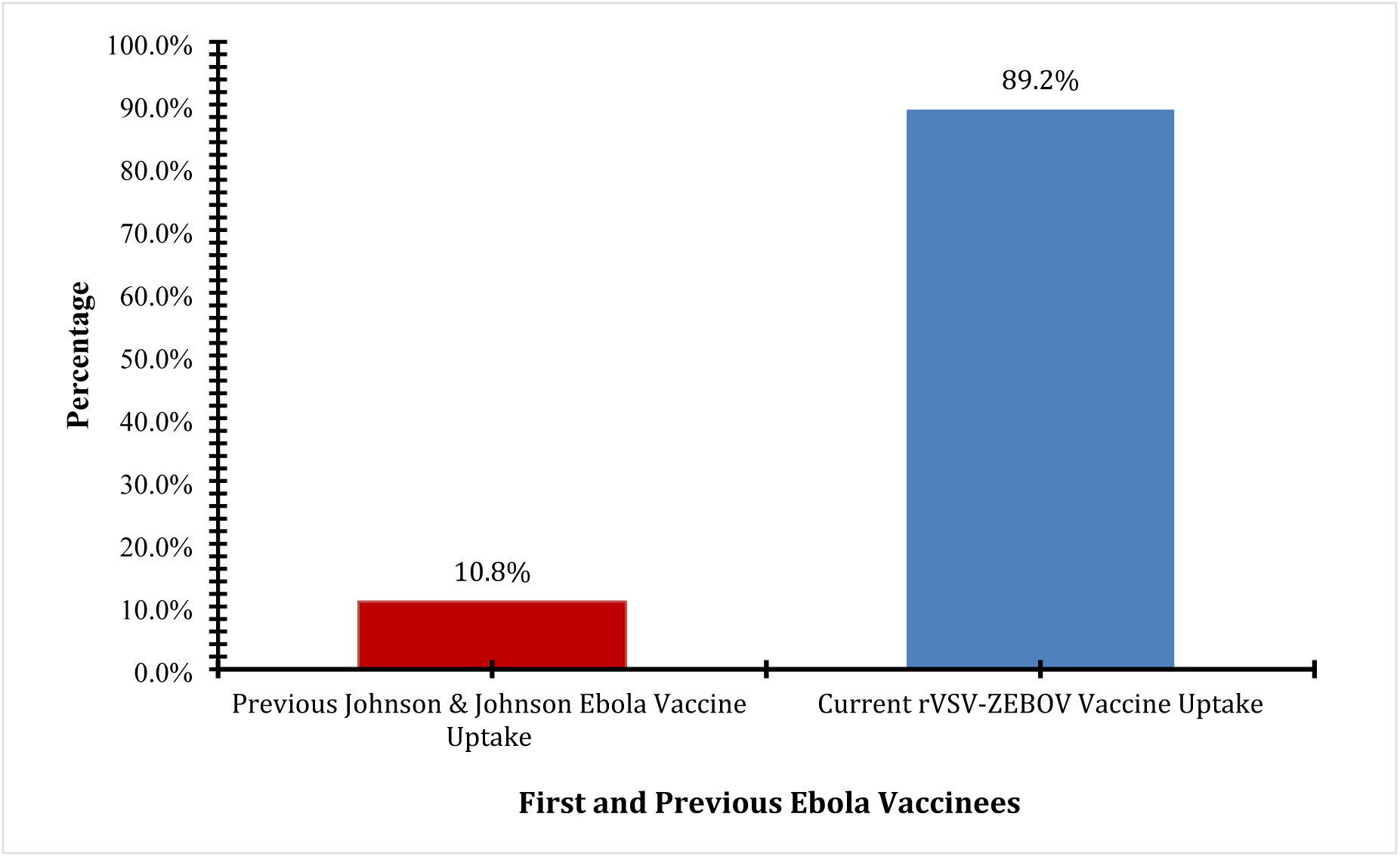
**Ebola Vaccination Status of Ebola Vaccine Vaccinees** Ebola Vaccination Status of Ebola vaccine vaccinees in Sierra Leone, December 2024. The graph illustrates a significant evolution in Ebola immunisation efforts within Sierra Leone, highlighting a clear divergence between the historical uptake of the Johnson & Johnson (Ad26.ZEBO/MVA-BN-Filo) two-dose regimen and the current trajectory of the rVSV-ZEBOV vaccine.

**Figure 2:**
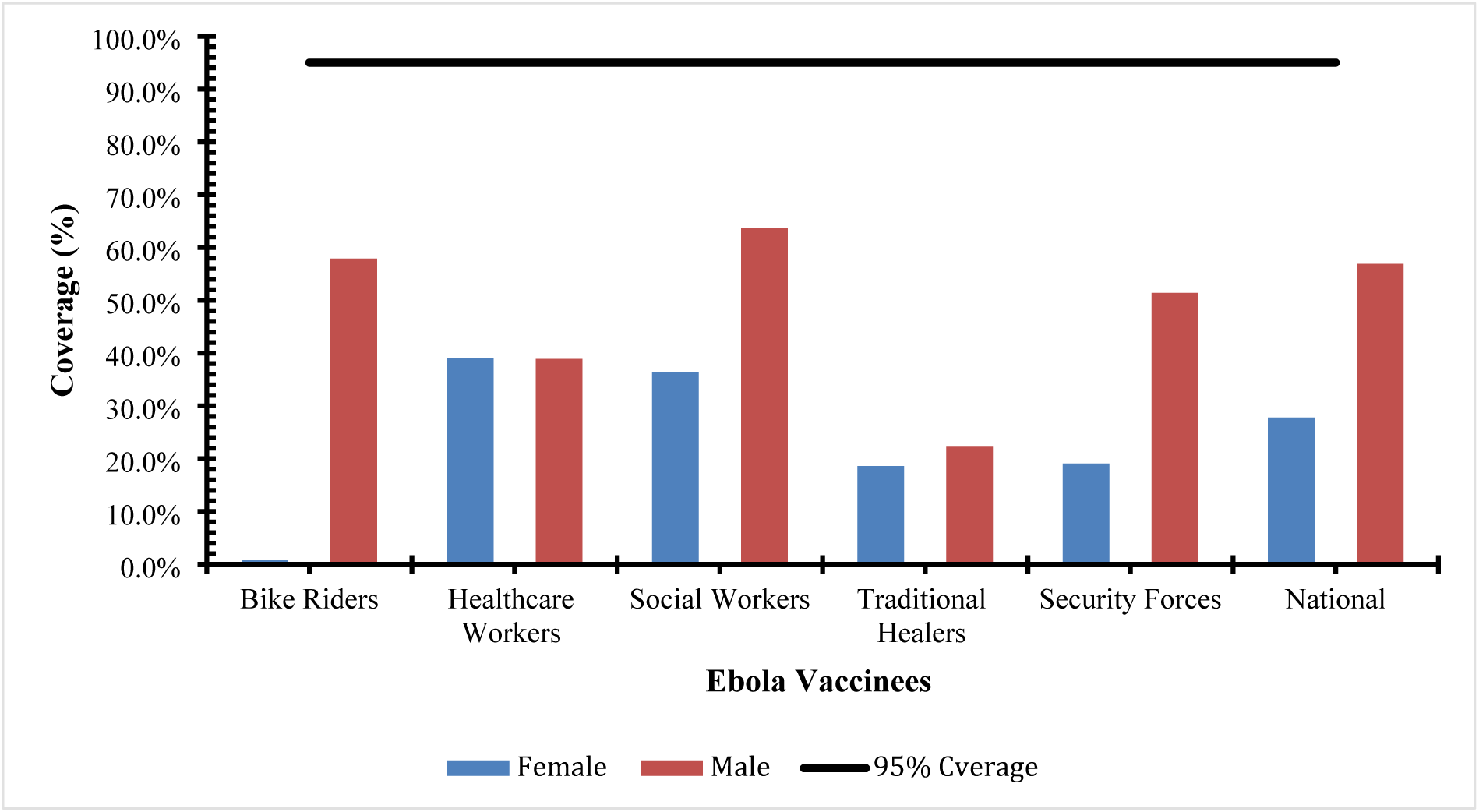
**Ebola vaccination coverage of Recipients by Districts in Sierra Leone Ebola vaccination coverage of Recipients by Districts in Sierra Leone** Ebola vaccination coverage of Ebola vaccine vaccinees in Sierra Leone, December 2024. This graph illustrates the cumulative percentages of Ebola vaccine vaccinees across the different target groups in Sierra Leone. Vaccination coverage for each target group was calculated as the total number of Ebola vaccine vaccinees during the Ebola vaccination campaign and divided by the estimated target population within the respective target groups, multiplied by 100.

### Sociodemographic Characteristics Associated with Ebola Vaccination Status

We used logistic regression analysis to determine the association between Ebola vaccination status and the sociodemographic characteristics of Ebola vaccine vaccinees. Individuals belonging to the age group 35 - 44 years had decreased odds (COR: 0.776, AOR: 0.761, P-value: 0.004) of Ebola vaccine vaccination compared to those belonging to the age group 18-24 years. Additionally, we discovered that the odds for Ebola vaccine vaccination further decreased for older age groups; 45-54 years (COR = 0.561, P < 0.001; AOR = 0.552, P < 0.001), and ->55 years (COR = 0.640, P < 0.001; AOR = 0.635, P < 0.001). The association between individuals belonging to the age group 25 - 34 years and Ebola vaccine status was not statistically significant (AOR = 0.963, 95% CI: 0.770 - 1.205, P-value = 0.742).

Regarding sex, differences were evident in Ebola vaccination. Females had higher odds of being vaccinated compared to males, with a COR of 1.433 (95% CI: 1.286-1.596, P < 0.001) and an AOR of 1.507 (95% CI: 1.293-1.756, P < 0.001).

Also, regional variations significantly impacted vaccination status. Individuals from the north-western region had a COR of 23.424 (95% CI: 19.950-27.503, P < 0.001) and an AOR of 25.502 (95% CI: 21.409-30.376, P < 0.001), indicating a markedly higher likelihood of vaccination. Similarly, those from the northern region showed a COR of 23.078 (95% CI: 19.259-27.655, P < 0.001) and an AOR of 39.539 (95% CI: 31.615-49.449, P < 0.001). The southern region also demonstrated high odds of vaccination COR of 27.576 (95% CI: 23.377-32.529, P < 0.001) AOR: 37.521 (95% CI: 30.886-45.581, P < 0.001), while the western region exhibited the highest odds with a COR of 43.293 (95% CI: 35.339-53.036, P < 0.001) and an AOR of 48.632 (95% CI: 38.317-61.723, P < 0.001).

However, marital status did not significantly affect vaccination status. Single individuals had a COR of 1.114 (95% CI: 0.554-2.241, P = 0.761) and an AOR of 0.888 (95% CI: 0.361-2.183, P = 0.795), while widowed or divorced individuals showed no significant differences compared to married individuals.

In terms of educational attainment, a significant association was seen with vaccination status. Individuals with primary education had a COR of 3.267 (95% CI: 2.692-3.964, P < 0.001) and an AOR of 1.437 (95% CI: 1.118-1.847, P = 0.005). Those with secondary education showed a COR of 0.869 (95% CI: 0.785-0.962, P = 0.007) and an AOR of 1.182 (95% CI: 1.022-1.368, P = 0.025). Individuals with tertiary education exhibited a COR of 2.405 (95% CI: 1.840-3.144, P < 0.001) and an AOR of 1.900 (95% CI: 1.371-2.633, P < 0.001). Also, occupation influenced vaccination status, particularly among healthcare workers. The odds of vaccination for healthcare workers were significantly lower, with a COR of 0.231 (95% CI: 0.176-0.302, P < 0.001) and an AOR of 0.684 (95% CI: 0.496-0.943, P = 0.021). In contrast, motorcyclists had an increased likelihood of being vaccinated (AOR: 2.075, 95% CI: 1.288-3.341, P = 0.003), while traditional healers served as the reference group.

**Table 3:**
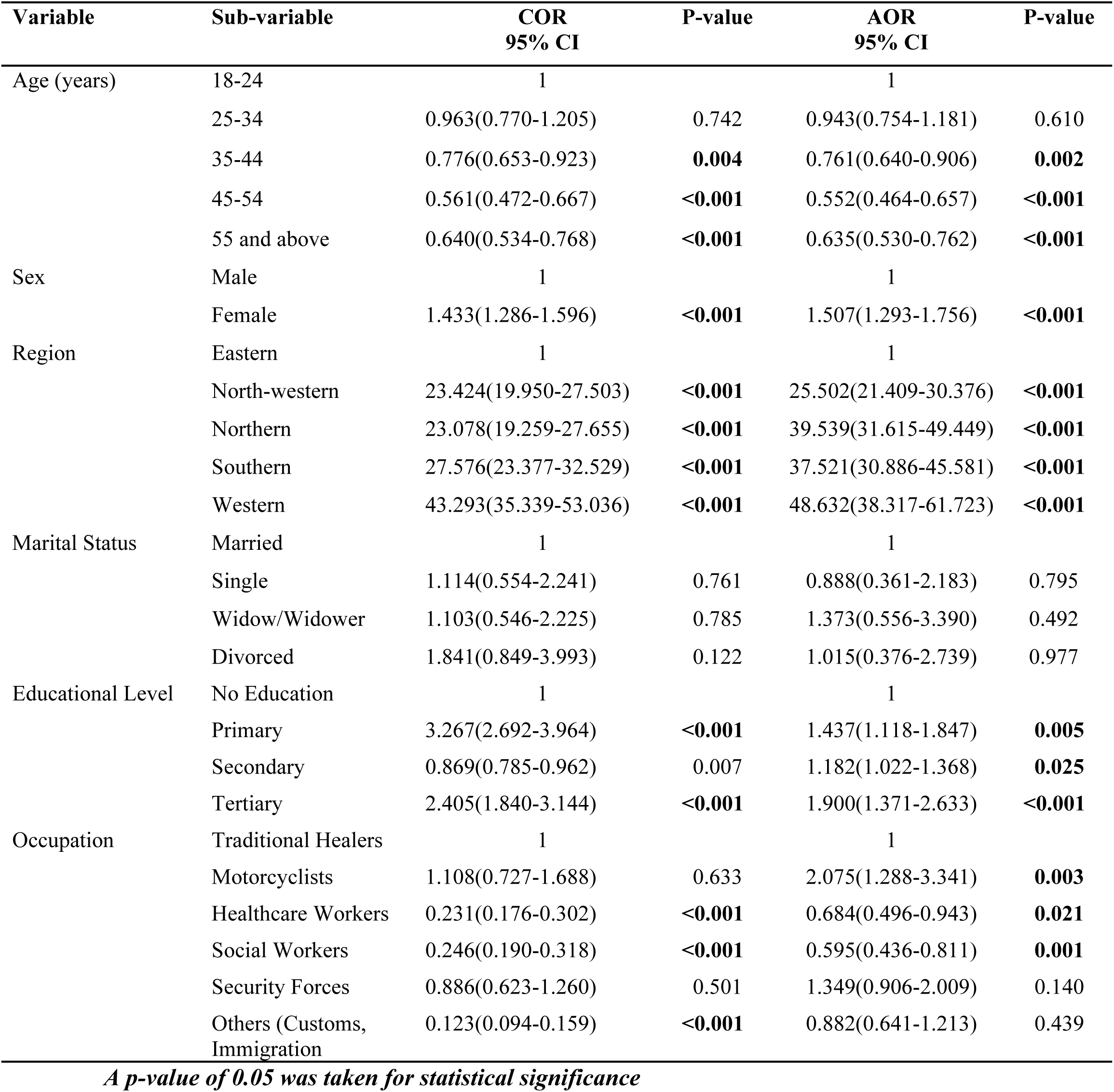
Bivariate and Multivariate Logistic Regression analysis showing the association of Ebola Vaccine Status among Ebola vaccine recipients in Sierra Leone, December 2024 (N=17454)

### Ebola vaccination coverage of Recipients by Districts in Sierra Leone

A coverage of 17,454 (84.64%) was achieved among the targeted population for the ERVEBO Ebola vaccination. Among the Ebola vaccinees, the majority, 11,729 (56.90%), were males. The majority of the targeted groups for the Ebola vaccination did not reach the 95% coverage set, except for social workers, 1,816 (100%). Additional information can be seen in Figure 3.

### Evolution of ERVEBO Ebola Preventive Vaccination Strategies in Sierra Leone Early Ebola Preventive Vaccination Campaign Efforts

The country introduced the Ebola vaccine using a targeted approach to prevent high-risk groups from contracting the virus in the future. Of the 16 districts in Sierra Leone, 8 districts that shared a border crossing with neighbouring Guinea were targeted for the vaccination. Healthcare workers were the main high-risk group who were vaccinated to prevent the spread of the virus in the country. The lessons learned from the previous vaccination in Sierra Leone show that individuals known to be high-risk groups in 8 out of 16 districts were vaccinated, leaving behind the other districts that have similar high-risk groups. In the bid to scale up vaccination and increase immunity against the Ebola virus, a nationwide vaccination strategy was developed and implemented to reach the high-risk population across the 16 districts. The target population included frontline healthcare workers, traditional healers, military officers, police officers, social workers, and commercial motorbike riders.

### Key Implementation Strategies: Ebola vaccination campaigns in Sierra Leone

Over the past decade, the Ministry of Health has positioned Sierra Leone’s health sector to implement various vaccination programs through the Expanded Program on Immunisation. Best practices for planning and implementing vaccination campaigns in Sierra Leone now sit deep in culturally sensitive community engagement and trust-building, not just for the Ebola vaccine but all other vaccines (15).

### Pre-vaccination Planning and Preparation

The rollout of the ERVEBO Vaccine in Sierra Leone was supported by Gavi, the Vaccine Alliance, the World Health Organisation (WHO), and partners from the International Coordinating Group on Vaccine Provision, aiming to safeguard the country’s healthcare workers while strengthening preparedness for future outbreaks (16). ERVEBO, an Ebola vaccine manufactured by Merck, is approved and licensed by the U.S. Federal Drug Administration (FDA) for individuals aged 12 months and older as a single-dose administration (17). Before the rollout of the Ebola vaccine in the country, the Ministry of Health, through the Pharmacy Board of Sierra Leone, which is the National Drug Regulatory Authority of Sierra Leone, approved the vaccine for use in the country after scientific data of the ERVEBO Ebola vaccine were made available by Merck.

Before the preventive Ebola vaccination started, the WHO Readiness Assessment Tool was adopted at the national and district levels. Three rounds of readiness assessments were conducted at these levels. The assessments took place periodically at intervals of four weeks, two weeks, and one week before the implementation. The readiness assessment tool consisted of six components: coordination, leadership and finance, service delivery, logistics, cold chain, waste management, monitoring and evaluation, and demand creation (18). Partners performed the national readiness assessment, while the national Expanded Programme on Immunisation staff carried out the district assessment. A total of twenty staff members were deployed across all districts to conduct the district-level readiness assessments for one day.

### The Effectiveness of communication and community engagement strategies in promoting vaccine acceptance and uptake

The findings show that early and accurate communication was central to the successful implementation of the preventive Ebola vaccination campaign. Social mobilisation activities, supported by IEC materials and mass media, helped prepare communities and improve understanding of the ERVEBO vaccine. Training of Trainers and stakeholder engagement facilitated effective information cascading, while targeted sensitisation on vaccine benefits, side effects, and prioritisation of high-risk groups contributed to increased acceptance of the vaccination programme.

### Logistics and Supply Chain Management of the Ebola Vaccine

Qualitative findings indicate that the ERVEBO Ebola vaccine was received, stored, and distributed in accordance with established cold chain and logistics protocols. The vaccine was cleared at the national level, stored under manufacturer-recommended conditions, and distributed using an approved distribution matrix developed through coordination between logistics and monitoring teams. Vaccines were centrally thawed and distributed to districts at recommended temperatures, with mixed cold chain equipment used to reach hard-to-reach areas. Daily last-mile distribution was conducted alongside vaccination teams, ensuring timely availability of vaccines throughout the campaign.

### Vaccination Team Composition and Training on the Preventive Ebola

The findings show that the vaccination campaign was implemented using a structured team deployment model, with each district deploying one static and two mobile teams coordinated by the District Medical Officer and supported by supervisors and partners. Comprehensive training was conducted at national, regional, and district levels prior to implementation, including Training of Trainers and targeted sessions for vaccinators, data officers, and supervisors. The use of standardised training materials and both paper-based and electronic data systems supported effective service delivery, accurate reporting, and timely data synchronisation throughout the campaign.

### Implementation of the ERVEBO Ebola Vaccination Campaign

The findings show that, despite delays in funding that prevented a phased rollout, the ERVEBO Ebola vaccination campaign was successfully implemented nationwide following a consensus decision by stakeholders. All districts conducted the campaign simultaneously, supported by strong coordination through a national situation room that enabled timely resolution of operational challenges. Continuous supervision by national, regional, district, and partner teams, using electronic tools, strengthened oversight, while systematic recording of beneficiary information supported effective monitoring and accountability throughout the campaign.

## Discussion

The successful rollout of the ERVEBO Ebola vaccine in Sierra Leone marks a significant milestone in the country’s decade-long journey toward epidemic preparedness and response. The national campaign achieved a vaccination coverage of 84.64% among 20,645 targeted high-risk individuals (healthcare workers, bike riders, security forces), underscoring the effectiveness of coordinated national planning, partner engagement, and trust-centred community participation. These results reflect substantial progress compared to the fragmented outbreak response during the 2013-2016 Ebola epidemic, when Sierra Leone’s health system faced systemic collapse and lacked any immunisation capacity for Ebola virus disease (19) (2).

### Public Health Impact and Vaccination Coverage Achievements

The high vaccination coverage observed aligns with findings from the WHO and CDC (2025) reporting increased vaccine acceptance among frontline workers in Sierra Leone following the introduction of preventive Ebola vaccination (16,20). This achievement demonstrates the feasibility of large-scale deployment of the single-dose rVSV-ZEBOV (ERVEBO) vaccine in low-resource environments (21). Comparable outcomes were observed in Guinea and the Democratic Republic of Congo, where ring vaccination achieved over 90% coverage among contacts and frontline personnel (22) (23). The data further revealed that healthcare workers (35%), followed by security forces and motorcyclists, represented the most vaccinated occupational categories. This aligns with the WHO Ebola vaccine framework, which prioritises individuals with the highest risk of exposure (WHO, 2021). The equitable coverage across most of the sixteen districts, particularly Bo, Port Loko, Kailahun, Kambia, and Koinadugu, achieving ≥100% coverage, highlights strong local coordination and vaccine distribution networks, enabled by the Expanded Programme on Immunisation (EPI) and partners such as Gavi and UNICEF (24) (25).

### Determinants of Vaccination Uptake

Multivariate analysis identified gender, region, education, and occupation as significant determinants of vaccination status. Females had 1.5 times higher odds of vaccination compared to males, echoing studies that link women’s greater trust in health workers and community networks to vaccine acceptance (26). Educational attainment strongly influenced uptake; participants with tertiary education were nearly twice as likely to be vaccinated as those with no education. This suggests that literacy and access to reliable health information remain central to vaccine acceptance (27). Regional variation in vaccine coverage also reflected disparities in logistics capacity, health infrastructure, and social mobilisation intensity. Districts in the western and southern regions recorded the highest odds of vaccination, which correlates with their superior cold-chain systems and health service readiness, as noted in WHO’s Service Availability and Readiness Assessment (15). Conversely, slightly lower coverage in Bombali and Kenema may indicate logistical bottlenecks or socio-cultural hesitancy rooted in prior outbreak trauma (28).

### Effectiveness of Communication and Community Engagement

Community engagement proved pivotal in driving vaccine uptake and countering misinformation. The government and partners implemented pre-vaccination social mobilisation using radio dramas, IEC materials, and faith-based advocacy methods proven effective in Ebola and COVID-19 vaccination campaigns across Africa (29,30). In Sierra Leone, trusted community figures such as chiefs and religious leaders acted as “social proof agents,” bridging the gap between medical authorities and communities, consistent with the “Four R’s” of engagement: reciprocity, relatability, relationships, and respect (30).

This culturally grounded communication strategy was essential for mitigating the effects of rumour proliferation and vaccine scepticism. Similar findings have been reported in studies exploring vaccine anxieties in Kono District (31) and the Democratic Republic of Congo (32), confirming that social cohesion and community endorsement are stronger determinants of uptake than availability alone.

### Cold Chain and Logistics Strengthening

The study’s findings on cold-chain functionality underscore the success of Sierra Leone’s supply chain strengthening efforts. Vaccines were maintained at 85°C until thawing and distributed using solar-powered refrigerators and Arktek devices, allowing temperature integrity even in hard-to-reach areas (Unicef, 2025). This mirrors best practices documented by Scott et al., who emphasise the use of renewable energy and last-mile delivery innovations in resource-constrained settings (33). The zero rate of Adverse Events Following Immunisation (AEFI) reported in this study further validates the robustness of vaccine handling and monitoring protocols. Moreover, logistical performance benefited from the integration of electronic supervision tools (Kobo Collect) and real-time data synchronisation. These digital approaches reflect WHO’s digital health transformation goals and align with lessons from the COVID-19 vaccine deployment, where data-driven monitoring enhanced accountability and timeliness (34).

### Lessons for Future Epidemic Preparedness

The ten-year trajectory of Ebola prevention in Sierra Leone illustrates the transformation of crisis into resilience. From the 2014 epidemic’s systemic collapse to the 2024 nationwide preventive vaccination, the country has built critical capabilities in risk communication, cold-chain infrastructure, and surveillance coordination. The incorporation of readiness assessments at national and district levels covering leadership, logistics, and waste management reflects a maturing health system now better equipped to handle emerging threats.

Key lessons for future implementation include the necessity of sustained investment in health workforce motivation, equitable access across all districts, and continued partnerships between public health agencies, academia, and communities. As Sierra Leone advances its preparedness framework, integrating Ebola vaccination into routine immunisation for high-risk occupational groups may offer cost-effective, long-term protection.

### Policy and Research Implications

From a health economics perspective, preventive vaccination represents a highly cost-effective intervention, especially when benchmarked against the economic losses and mortality incurred during the 2014-2016 outbreak. Economic modelling studies estimate that vaccination campaigns yielding >80% coverage can reduce outbreak costs by 40-60% and avert thousands of deaths (36). Future evaluations should incorporate cost-per-dose delivered and value-for-money metrics to optimise donor and government investments.

Additionally, the establishment of data-driven vaccine registries and digital logistics management systems will be vital for sustainability. Continuous community dialogue should remain central to overcoming hesitancy and maintaining confidence in health interventions.

## Conclusion

This study confirms that Sierra Leone’s Ebola vaccination efforts over the past decade have evolved from reactive outbreak management to proactive disease prevention. The country’s achievements in high coverage, safe cold-chain operations, and strong community engagement demonstrate that even resource-limited nations can attain vaccine equity through partnership, innovation, and trust. These lessons are not only vital for Ebola preparedness but also provide a model for future epidemic response in sub-Saharan Africa.

## Author Contributions

Conceptualisation: DMK, ENS, and NCK; manuscript acquisition: DMK, ENS, NMK, and NCK; Methodology: DMK, ENS, NMK, AKK, SL, MJ, and NCK; formal analysis: SL, AKK, NMK, and ENS; writing review and editing: JK, PBJ, DMK, HB, HC, NMK, AKK, ENS, NCK, MJ, EGS, and SL; Visualisation: DMK, SL, and ENS. All authors have read and agreed to the published version of the manuscript. All authors agree to be accountable for all aspects of the work in this article.

## Institutional Review Board Statement

The study was not considered human subject research; hence, IRB is not applicable.

## Informed Consent Statement

Not applicable

## Data Availability Statement

The data underlying this article were provided by and are the property of the Ministry of Health of Sierra Leone. The data will be shared on a reasonable request with the ministry’s permission.

## Data Availability

All relevant data are within the manuscript and its Supporting Information files

## Acknowledgement

This study was conducted by the Expanded Programme on Immunisation (EPI), Ministry of Health, Sierra Leone. The authors thank the healthcare workers and monitoring and evaluation officers. We are also grateful to our partners, CHAI, UNICEF, and WHO.

